# Understanding Purchasing Patterns of Alcoholic, Alcohol-Free, and Low-Alcohol Drinks: A Latent Profile Analysis

**DOI:** 10.64898/2025.12.03.25341542

**Authors:** Oscar Rousham, Abigail K. Stevely, John Holmes

## Abstract

**Background:** Alcohol-free and low-alcohol (no/lo) drinks (*≤*1.2% ABV) are increasingly popular in high-income countries. Their potential to reduce alcohol-related harm depends on who buys them, in what quantity, and their incorporation into overall drinking patterns. We aimed to (i) compare characteristics of purchases containing only no/lo drinks, only alcoholic drinks, or both, over time between 2018 and 2023; (ii) identify subgroups with distinct purchasing patterns in 2023; and (iii) describe purchasing and soiodemographic differences between these subgroups.

**Design:** Latent profile analysis of cross-sectional household purchasing data.

**Setting:** Great Britain, 2018 and 2023.

**Participants:** Nationally representative samples of 30,401 (2018) and 28,254 (2023) households. 4,975 households who purchased no/lo drinks in 2023 were included in the latent profile analysis.

**Measurements:** Data included off-trade (i.e. shop) purchases categorised into: no/lo-only; alcohol-only; or no/lo alongside alcohol. Household characteristics were frequency of each purchase type; standard servings of no/lo drinks per adult; alcohol risk levels based on weekly units of alcohol purchased per adult in household (non-drinker: 0 units; low-risk: *≤*14 units; increasing risk: >14 *≤*35 units; high-risk: >35 units [1 unit = 8g alcohol]); household age; social class; region; and ethnicity.

**Findings:** From 2018 to 2023, the proportion of purchases that were alcohol-only fell from 97% (95% CI: 97%-97%) to 95% (CI: 95%-95%) while no/lo-only purchases rose from 1.4% (CI: 1.3%-1.4%) to 2.7% (CI: 2.7%-2.8%) and no/lo alongside alcohol purchases rose from 1.2% (CI: 1.2%-1.2%) to 1.9% (1.9%-2.0%). In 2023, no/lo-only purchases contained fewer servings (median = 6.9) than no/lo alongside alcohol purchases (median = 6.5 plus 24.5 alcohol units) and alcohol-only purchases (median = 24.6 units). No/lo-only purchases occurred earlier in the week, no/lo alongside alcohol purchases peaked on Fridays and Saturdays.

Latent profile analysis identified three classes: (i) *no/lo triers* (53%) averaged 2.1 no/lo servings per adult annually with 95% purchasing no or low-risk levels of alcohol. (ii) *Occasional purchasers* (34%) averaged 7.5 servings with 20% purchasing alcohol at increasing or high-risk levels. (iii) *Dual purchasers* (13%) averaged 37.8 servings with 39% purchasing alcohol at increasing or high-risk levels. *Dual purchasers* and *occasional purchasers* were older than *no/lo triers* (60% [p<0.001], 54% [p=0.010], and 49% 55 years respectively). *Dual* and *occasional purchasers* were more often white than *no/lo triers* (97% [p = 0.014], 97% [p=0.0074], 94% respectively).

**Conclusions:** In Great Britain, most households purchasing no/lo drinks do so infrequently and purchase alcohol at low-risk levels; however, a smaller group of older, higher-risk households purchase no/lo drinks more frequently.

## BACKGROUND

Alcohol is a major public health problem, accounting for 5.3% of deaths globally ^1^, and is the biggest risk factor for death, illhealth, and disability among 15 - 49 year-olds in England ^2^. Alcohol-related harms are unequally distributed, with lower socioeconomic groups experiencing more harm from less alcohol consumption (Office for National Statistics, 2023). Alcohol also imposes a substantial financial burden, costing the National Health Service in England an estimated £4.9bn each year ^3^.

In the UK, alcohol-free and low-alcohol (no/lo) drinks, defined as resembling alcoholic beverages but containing *≤*1.2% alcohol by volume (ABV), have seen a rapid increase in popularity since 2018. In 2023, they accounted for 1.3% of sales, up from 0.9% in 2020^4^. The current UK government has continued the previous governments commitment to supporting industry to increase no/lo drinks availability as a strategy to reduce alcohol consumption and related harms ^5,6^. However, there are concerns that the increased popularity of no/lo drinks may increase alcohol consumption by normalising drinking in new contexts ^7^ or enabling surrogate marketing; that is, advertising no/lo drinks which share branding with their alcoholic counterpart in order to circumvent advertising restrictions ^8^.

To reduce alcohol harms, no/lo drinks must be consumed as a substitute for, rather than in addition to, alcohol and at a scale which meaningfully reduces alcohol consumption ^9^. To maximise public health benefits and reduce inequalities, this substitution should occur among heavier drinkers, particularly those of lower socioeconomic position ^10^. Evaluating the public health impacts of no/lo drinks therefore requires an understanding of how they are incorporated into drinking patterns, who consumes them, and in what quantities. In effect, no/lo beverage drinking patterns. As with alcohol, no/lo drinking patterns are likely to vary across intersecting sociodemographic characteristics ^11,12.^

Research on no/lo beverage drinking patterns remains limited. Self-report data indicate that these drinks are sometimes consumed alone, for example, when driving or before an early start ^9,^ and that certain sociodemographic groups are more likely to consume them alongside alcohol ^13^. Anecdotal evidence also highlights zebra striping, occasions where drinkers alternate between alcoholic and non-alcoholic drinks to moderate intake ^14^. However, no previous studies have used purchasing data to examine how no/lo beverages are integrated into drinking patterns.

There is emerging evidence that men, heavier drinkers, and individuals of higher socioeconomic position (SEP) are more likely to consume or purchase no/lo drinks ^13,15,16.^ Findings regarding age are inconsistent, however, with different studies finding that no/lo consumers are younger ^17^, older ^13^, or showing no evidence for age differences ^16^. These studies are limited by treating no/lo consumption as a binary behaviour (ever vs never consumed) rather than examining the quantity consumed. They also treat no/lo consumers as a homogenous group, overlooking the possibility of distinct subgroups of consumers.

There is a need for a more comprehensive understanding of how no/lo beverages are being incorporated into alcohol consumption patterns and for research which captures the heterogeneity in who consumes no/lo beverages and how much they consume. Given the rapid increase in popularity of no/lo drinks since 2018, there is a need to understand how consumption patterns of no/lo beverages have changed over time.

### Aims and Research Questions

Whilst not identical (see Limitations), purchasing data offer a large-scale, recall-bias-free insight into consumption. Using household purchasing data for alcoholic and no/lo drinks purchased in the off-trade (i.e. supermarkets and other stores) we aim to: (i) understand the characteristics of no/lo drinks purchases and how this has changed over time, (ii) identify the different subgroups of no/lo consumers and (iii) describe their behavioural and sociodemographic characteristics.

#### RQ1

How do purchase-level characteristics of three purchase types (solely no/lo beverages, solely standard alcoholic beverages, and no/lo alongside standard alcoholic beverages) compare in terms of timing, volume, and cost? How has this changed between 2018 and 2023?

#### RQ2

Can we identify subgroups of households with distinct purchasing patterns according to their annual purchase frequency of these three purchase types in 2023?

#### RQ3

How do households following different purchasing patterns differ in their annual standard alcoholic and no/lo beverage purchasing and their sociodemographic characteristics?

## METHODS

The data analysis plan was pre-registered on the Open Science Framework on 10th February 2025: https://osf.io/9p7hj/

### Data

We used data from Kantars World Panel dataset (KWP) from 2018 and 2023 (the earliest and latest years available to us at the time of analysis). KWP tracks daily off-trade purchases for a continuous household purchasing panel of 30,000 nationally representative households in Great Britain. A member of each household scans barcodes of food and drink products brought into the home. We have access to data on alcoholic and no/lo beverage purchases and household sociodemographics. Households are recruited by stratified quota sampling based on region and sociodemographics (e.g. household size, children, age of main shopper) and included in the dataset if they meet quality standards including minimum thresholds for recording, purchase volume, and spend. Socioeconomic group is part of the weightings used to ensure a representative population, which we use in RQ1. Households that leave the sample are replaced on a continuous basis to maintain a representative sample. Each year is divided into 13 four-week periods.

For descriptive statistics (RQ1), we included all purchases made in 2018 (n = 497,821 purchases by 30,401 households) and 2023 (n = 379,205; 28,254). For latent profile analysis (RQ2), we included households present in the dataset for at least three four-week periods and with at least one no/lo drinks purchase in 2023 as our focus was on purchasing patterns that included no/lo beverages (after imputation n = 4,975). Households without no/lo drinks purchases were reintroduced after model fitting for sociodemographic comparisons (RQ3).

### Measures

A purchasing occasion was defined as all purchases by a household on a single day and classified into three types: (1) no/lo drinks only, (2) alcohol only, or (3) no/lo drinks alongside alcohol. At the level of the purchasing occasion, purchase timing was measured as the proportion of daily purchases by type for each day of the week between 2018 and 2023. Purchase frequency (in 2018 and 2023) was the annual number of occasions across the whole sample for each purchase type. Alcohol volume in units (1 unit = 10ml pure alcohol), and no/lo volume in servings, defined by drink type (Beer: 330ml; Wine: 175ml; Cider: 500ml; Spirits: 50ml; RTDs: 250ml) ^18,^ were measured for each purchasing occasion in 2018 and 2023.

At the household level, three behavioural measures were captured: annual purchase frequency by type, alcohol risk category based on the mean units of alcohol purchased per week across the year and number of adults in the household (non-drinker: 0 units; low-risk: *≤*14 units per adult per week; increasing risk: >14 *≤*35 units; high-risk: >35 units), and annual no/lo servings per adult.

Household sociodemographic measures included region (England, Wales, Scotland); social class, recorded by Kantar Worldpanel according to the National Readership Survey Classifications ^19^ and collapsed into AB (Higher or intermediate managerial, administrative or professional), C1 (junior managerial, administrative or professional), C2 (skilled manual workers), and DE (semi- or unskilled-manual workers, casual workers or unemployed) ; main shopper ethnicity (Asian, Black, Mixed, White, Other) and age (<28, 2834, 3544, 4554, 5564, 65+); and number of adults (calculated from the data available in KWP using household size minus number of children).

## ANALYSIS

### Descriptive Statistics (RQ1)

At the purchase level, for each purchase type, we calculated the number of purchases and the median and interquartile range of both cost and volume of alcohol and no/lo drinks across all purchases of that type for 2018 and 2023. For purchase timing, the mean proportion of purchases that were on each day across all weeks and 95% confidence intervals calculated using Pearson’s chi-squared test. All descriptive statistics apart from number of purchases were weighted using KWP panel weightings.

### Latent Profile Analysis (RQ2-3)

Latent profile analysis (LPA) is a clustering method used to split heterogeneous populations into smaller more homogenous groups, called classes, using numerical variables. We used LPA to categorise no/lo-purchasing households in 2023 according to their purchase frequency across our three purchase types. Models were fit iteratively: starting with one class, additional classes where retained where model fit improved according to BIC (Bayesian information criteria), AIC (Akaike information criteria) and the bootstrap likelihood ratio test. Models producing latent classes containing <5% of participants were excluded, as these typically reflect unsorted cases rather than meaningful subgroups. Model entropy was reported but not used to choose the final model. All models allowed class-specific variance (i.e. spread of purchase frequencies about the mean) and covariance (i.e. relationship between the frequency of each of the three purchase types) structures. For subsequent analyses, households were assigned to the class with the highest posterior probability of membership.

To characterise each latent class, we calculated the mean annual servings of no/lo beverages per adult and the proportion of households in each alcohol risk category. Non-purchasing households were split into alcohol-only purchasers and nonalcohol purchasers and described in the same way. As our aim was descriptive rather than causal, sociodemographic characteristics of each class were summarised and compared using pairwise statistical tests, treating the largest class as the reference group.

We were unable to weight the LPA analyses due to software constraints.

Ethics approval was obtained from the University of Sheffield, application number: 052135. Analyses were conducted in R ^20^. Mplus ^21^ via the Mplusautomation ^22^ and tidyLPA ^23^ packages were used to fit latent profile models.

### Missing Data

For each four-week period that a household was not included in the dataset, purchase type frequencies and the volume of alcoholic and no/lo drinks of each beverage category (beer, wine, etc) purchased were imputed using multiple imputation by chained equations (MICE). To capture seasonal variation, imputation was conducted separately for each four-week period before summing for annual frequency and volume.

Imputation introduces uncertainty in purchase frequency which affects both the optimal number of latent classes and household classification. To address this, we generated 100 imputed datasets, identified the optimal class number in each using the criteria above, and selected the most frequently occurring solution as final. A model with this number of classes was then fit to all 100 datasets, with households assigned according to posterior probability ^24^.

We also accounted for this uncertainty when calculating latent class characteristics: For household counts, proportions of non-drinkers, and alcohol risk categories, values were calculated within each imputation then averaged. For servings of no/lo drinks, the median for each class was calculated within each imputation then across imputations. Sociodemographic characteristics were based on the class households were most often assigned to across imputations.

To impute purchase frequency, we used auxiliary variables on purchasing behaviour (frequency and volume during observed periods) and sociodemographic characteristics (education, social class, region, ethnicity, age of main shopper). Volume was imputed after fitting latent profile models, allowing class membership to be included as an auxiliary variable. Missingness was assumed to be missing completely at random (MCAR) or at random (MAR).

### Changes from the published analysis plan

We made three deviations from the pre-registered analysis plan (https://osf.io/9p7hj/). First, household-level purchase frequencies by type were omitted due to space constraints and overlap with RQ2. As a result, the inclusion criteria for RQ1 could be expanded to all purchases in 2018 and 2023. Second, mean annual alcohol purchasing was replaced with alcohol risk levels, which we considered more interpretable. Third, purchase timing was calculated across 2018 - 2023 rather than 2023 alone to retain useful data.

## RESULTS

### Purchase Level Descriptive Statistics

In 2023 there were 361,708 alcohol-only purchases, 7,562 of no/lo alongside alcohol, and 9,935 no/lo-only (Table 1). Thus, 44% of unweighted no/lo purchases in Great Britain contained no alcohol, while 56% did.

**Table 1:** Purchase-level characteristics for three purchase types: alcohol only, no/lo drinks alongside alcohol, and no/lo drinks only.

|  |  | Alcohol only |  | No/lo drinks + alcohol |  | No/lo drinks only |  |
| --- | --- | --- | --- | --- | --- | --- | --- |
| Purchase-level characteristics |  | 2018 | 2023 | 2018 | 2023 | 2018 | 2023 |
| Number of purchases (% [95% CI]) |  | 458,349 (97 [97-97]) | 361,708 (95 [95-95]) | 5,948 (1.2 [1.2-1.2]) | 7,562 (1.9 [1.9-2.0]) | 6,524 (1.4 [1.3-1.4]) | 9,935 (2.7 [2.7-2.8]) |
| Median volume (IQR) | Alcohol (Units of alcohol) | 22.3 (36.4) | 24.6 (40.6) | 21.3 (40.2) | 24.5 (43.4) | – | – |
|  | No/lo drinks (Number of servings) | – | – | 6.3 (8.5) | 6.5 (8.3) | 7.0 (7.9) | 6.9 (9.2) |
| Median price (£) (IQR) |  | 11.7 (17.8) | 14.5 (22.0) | 16.9 (22.5) | 23.2 (29.6) | 3.94 (4.6) | 5.79 (7.56) |

From 2018 to 2023, alcohol-only purchases fell from 97% to 95% of total purchases, no/lo alongside alcohol rose from 1.2% to 1.9%, and no/lo-only saw the largest growth, from 1.4% to 2.7%. Over the same period, median alcohol volume rose from 22.3 to 24.6 units in alcohol-only purchases, and from 21.3 to 24.5 units in no/lo alongside alcohol purchases. Median no/lo drink volumes remained stable, from 6.3 to 6.5 servings when bought alongside alcohol and 7.0 to 6.9 servings when purchased alone. Purchases combining alcohol and no/lo drinks were the largest and most expensive, with alcohol volumes similar to alcohol-only purchases and no/lo volumes similar to no/lo-only purchases. In 2023, median prices were £23.20 for mixed purchases, £14.50 for alcohol-only, and £5.79 for no/lo-only. The proportion of purchases that were alcohol-only were lowest Mondays and Tuesdays and higher the rest of the week. Purchases of no/lo alongside alcohol peaked on Fridays and Saturdays whilst no/lo-only purchases showed the reverse pattern, lowest Thursdays and Fridays and highest Sunday to Wednesday.

### Latent Profile Analysis

A three-class model with variance and covariance differing by class was most frequently the best fit across imputations (68%; see table 2). These classes, defined by frequency of alcohol-only, alcohol alongside no/lo, and no/lo-only purchases (Table 2), were: *no/lo triers* (53% of no/lo households) who averaged just 2.1 no/lo servings in 2023 and mostly purchased alcohol at low risk levels (90%) or none (5.2%) (Figure 2); *Occasional purchasers* (34%) who averaged 7.5 no/lo servings and had higher rates of increasing (16%) and high-risk (4.4%) alcohol purchasing; And *dual purchasers* (16%) who were frequent purchasers averaging 37.8 servings and most likely to purchase alcohol at increasing (19%) or high-risk (10%) levels. *Dual purchasers* accounted for 58% of all no/lo servings purchased, compared to 28% for *occasional purchasers* and 14% for *no/lo triers* (Table 3).

**Figure 1:**
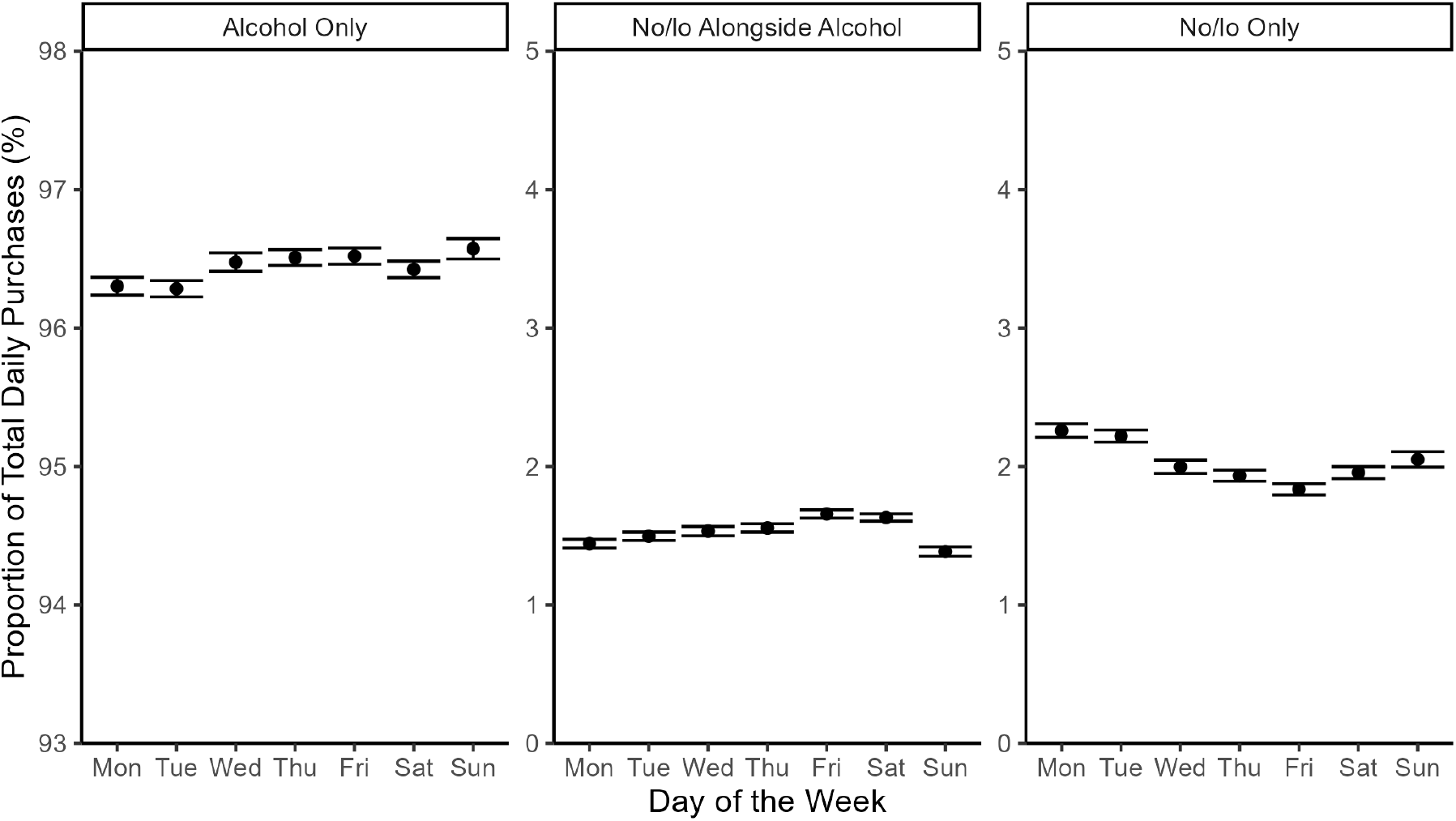
Proportion of total daily purchases of each purchase type by day of the week (%). Points represent the mean proportion; error bars represent the standard error around the mean.

**Table 2:** Model fit criteria.

| Table 2: Model fit criteria. |  |  |  |  |  |  |
| --- | --- | --- | --- | --- | --- | --- |
| Classes | AIC | BIC | Entropy | Class 1 (%) | Class 2 (%) | Class 3 (%) |
| 1 | -41,132 | -41,073 | 1.00 | 100 | – | – |
| 2 | -60,065 | -59,941 | 0.896 | 25.1 | 74.9 | – |
| 3 | -64,332 | -64,142 | 0.842 | 11.8 | 53.5 | 34.7 |
| 4 | Did not converge |  |  |  |  |  |

**Table 3:**
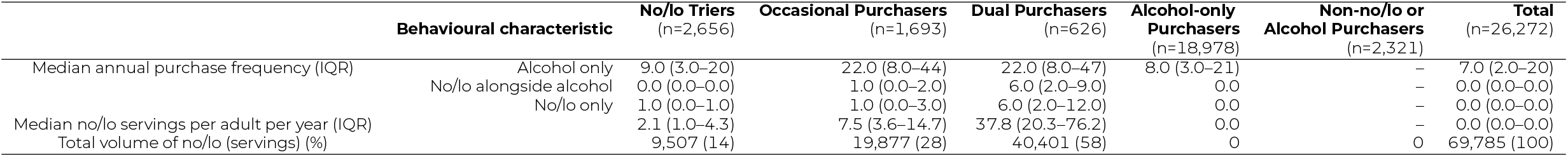
Behavioural characteristics by latent class.

**Figure 2:**
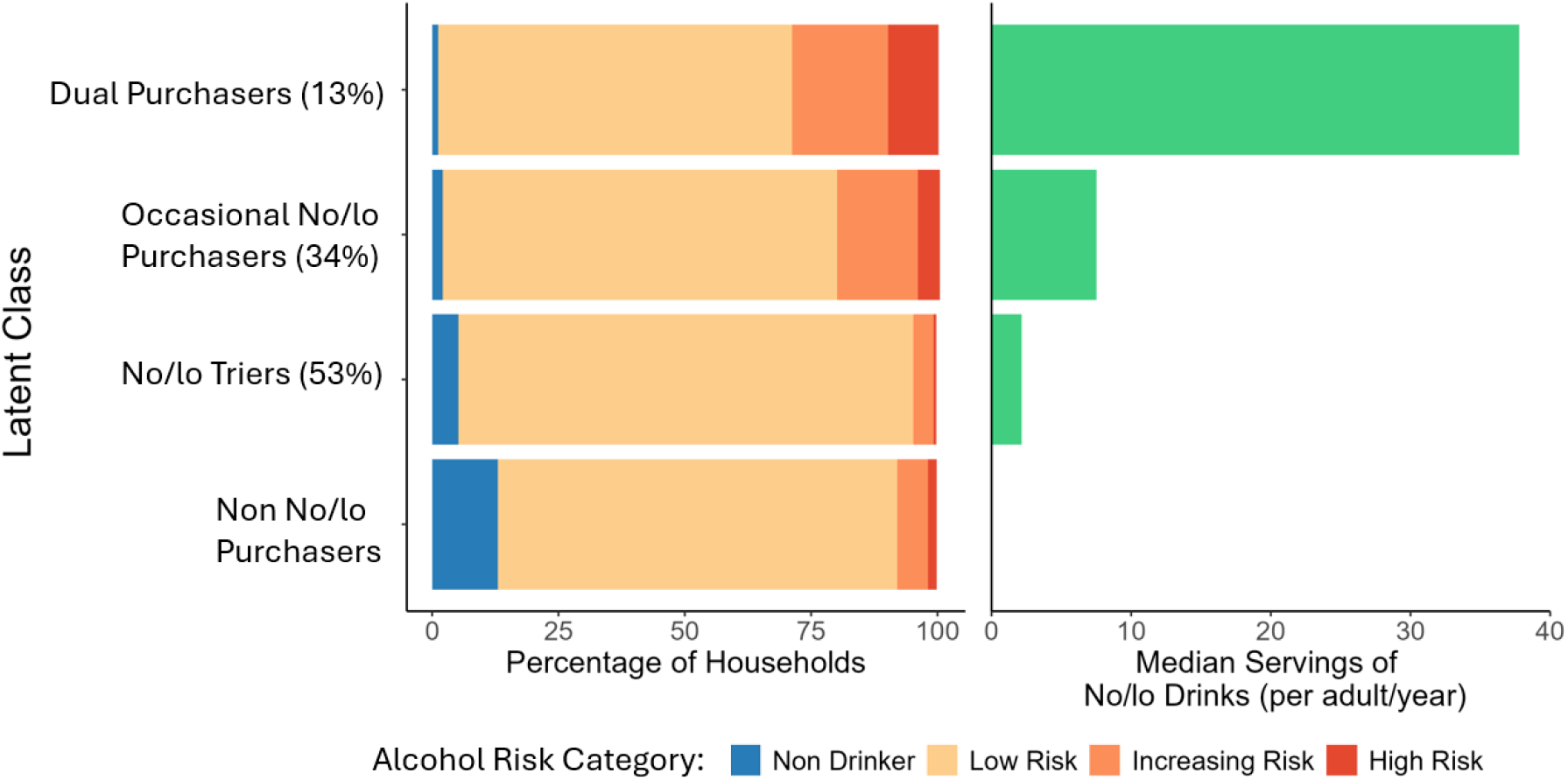
Proportion of each class which purchased alcohol at different risk levels in 2023 (left) and median servings of no/lo beverages in 2023 by class (right).

Regarding sociodemographic characteristics (Table 4), *occasional purchasers* were older than *no/lo triers* (54% vs 49% aged 55+ years) and *dual purchasers* older still (64%) *No/lo triers* resembled alcohol-only households in age distribution but were older than households purchasing neither alcohol nor no/lo.

**Table 4:**
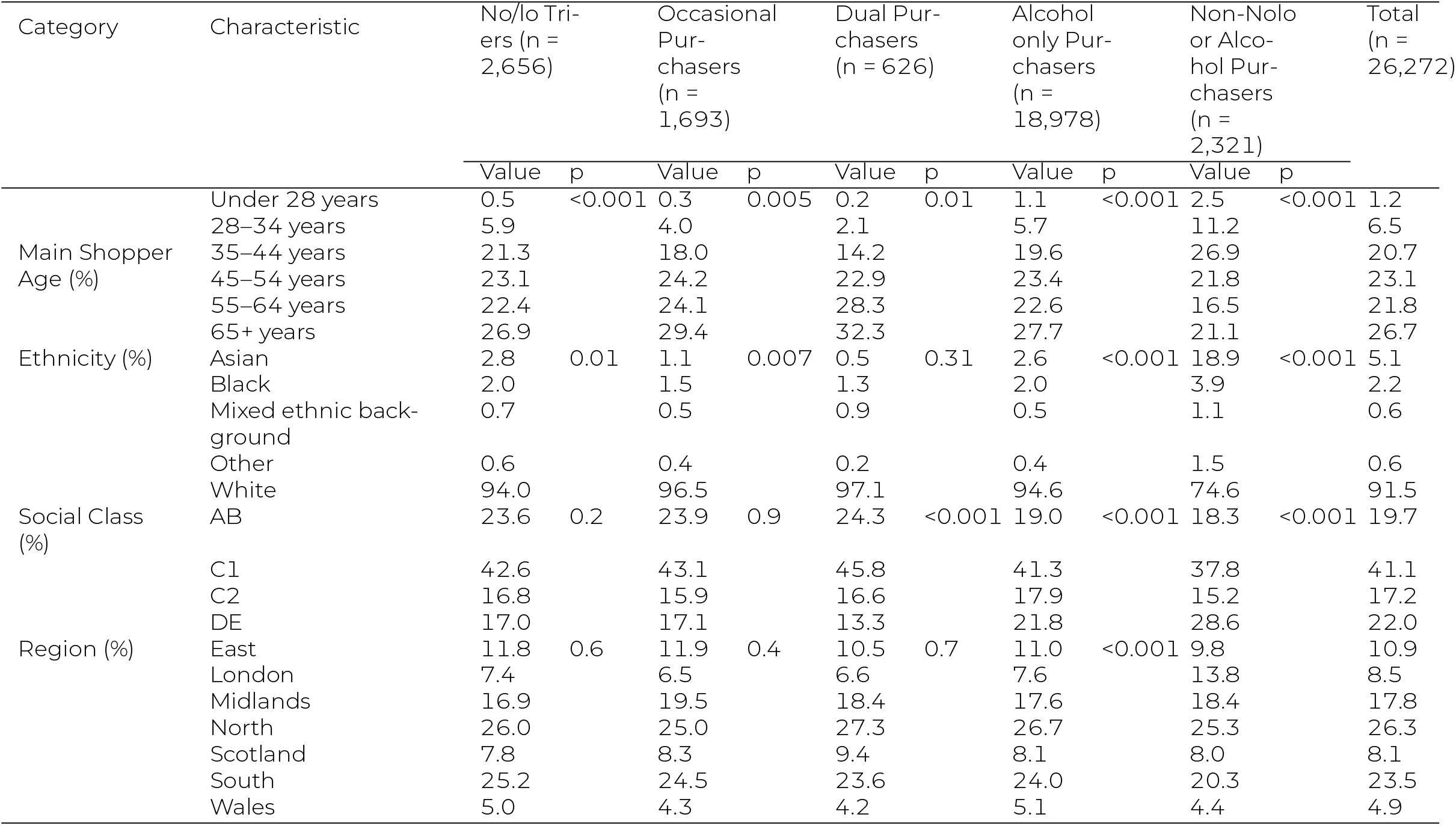
Sociodemographic characteristics by latent class.

*Dual* and *occasional purchasers* both had a significantly higher proportion of White individuals compared to no/lo triers (97% vs 94%). *No/lo triers* had a similar distribution of ethnicities to *alcohol-only purchasers* but *non-no/lo beverage, non-alcohol purchasers* had a significantly higher proportion of individuals from non-white backgrounds (24% vs 4%).

No differences in social class were observed between the three no/lo purchasing classes. However, all had a higher proportion from higher social classes than alcohol-only or non-alcohol, non-no/lo purchasers (24% AB vs 19% and 18%, respectively).

## DISCUSSION

### Key Findings

Around half of purchases containing no/lo drinks in Great Britain were solely no/lo beverages without any alcohol. These purchases tended to be smaller and more often on weekdays than those that contained alcohol. The remaining purchases containing no/lo drinks also contained alcoholic drinks and had the opposite temporal pattern, peaking later in the week on Fridays and Saturdays.

No/lo purchasers can be divided into three groups: *No/lo triers* (53%), *Occasional purchasers* (34%), and *dual purchasers* (13%). *No/lo triers* tend to be purchase alcohol at lower risk levels, make very few purchases of no/lo drinks and are similar to alcohol-only purchasers in terms of age, ethnicity and region. *Occasional purchasers* tend to purchase alcohol at increasing risk levels, purchase no/lo drinks at an intermediate rate and are older than *no/lo triers. Dual purchasers* tend to purchase alcohol at higher risk levels, purchase no/lo drinks regularly and are older than *occasional purchasers*.

### Strengths and Limitations

This study has several strengths. We used purchasing data which is free from recall bias and allows for granular analyses at the level of the purchasing occasion, although households may still under-report their purchases for other reasons. Applying latent profile analysis to these data captured heterogeneity in who purchases no/lo beverages and how much they purchase, overcoming limitations of previous analyses which treat no/lo consumption as a binary outcome by a homogenous group. Multiple imputation enabled the inclusion of a larger sample in the latent profile analysis, reducing the risk of bias from missing data. By using many imputations and running latent profile models on each dataset, we also accounted for the uncertainty introduced by the imputation process.

However, there are some limitations to our study. First, data were at the household level and based on purchasing, meaning we cannot determine how the consumption of purchases was distributed across households members, or the characteristics of the households members who consumed them. Second, our data only covers off-trade purchases, excluding those made outside the home, although the off-trade accounted for 73% of total alcohol purchases in 2023 (BBPA, 2023). Further, purchasing is not identical to consumption as products might not be consumed immediately, particularly for multipacks and large bottles of spirits. However, the datasets size and longitudinal coverage of both purchasing and non-purchasing periods helps moderate this.

### Interpretation and Comparison with Existing Literature

While off-trade purchases are not necessarily consumed immediately, the different volume and temporal patterns of each purchase type suggest the existence of distinct no/lo drinking occasions. Purchases of solely no/lo drinks earlier in the week may indicate either consumption on typically non-drinking days or full substitution for alcohol. This aligns with findings that people often choose no/lo drinks when they need to wake up early for work or other commitments ^9^.

The demographics of the three identified classes align with previous research. In line with Perman-Howe et al ^13^ and Clarke et al ^16^, we found no/lo purchasers were more likely to be from higher social classes. Our results suggest that previous mixed findings on the association of age with no/lo purchasing or consumption are not contradictory but reflect distinct groups of no/lo consumers: We found that one class of no/lo consumers had a similar age profile to non-no/lo drinkers, whilst our other two identified classes were older.

Interestingly, we did not identify a class of households which purchased solely no/lo beverages, suggesting that this purchasing behaviour is rare. However, given that our data is at the household level, it is possible that there are individuals who consume solely no/lo drinks but live with alcohol drinkers. This is in accordance with existing research showing that no/lo drinks consumption is more likely in alcohol drinkers than in non-drinkers ^13,16^.

### Implications of Findings

In 2023, over half of no/lo drinking households were classified into the *no/lo triers* class, averaging just one purchase of no/lo drinks in 2023. Even if members of this class were consuming no/lo drinks as a substitute for alcohol, the scale of this substitution is unlikely to reduce alcohol harms. This is especially the case given that 95% of households in this class purchased no alcohol or purchased alcohol at low-risk levels. A further 34% of households were categorised into the *occasional purchasers* class, purchasing on average just 7.5 servings of no/lo drinks per adult in 2023. This level of purchasing, equivalent to roughly one serving every seven weeks, also has little potential to reduce alcohol consumption to a degree that would reduce alcohol-related harms.

This limited potential of no/lo drinks to reduce alcohol-related harm for most no/lo purchasing households should be weighed against concerns that the increased availability of no/lo drinks may normalise alcohol consumption in new contexts ^7^ and enable surrogate marketing by the alcohol industry ^8^. It should be noted, however, that no/lo drinks are continuing to grow in popularity and patterns and volumes of no/lo purchasing may change as the market grows.

In contrast, whilst the *dual purchasers* class contained only 13% of no/lo purchasing households they accounted for 58% of no/lo purchases by number of servings in 2023. This class also contained the highest proportion of households purchasing alcohol at high-risk or increasing-risk levels. This purchasing could reduce alcohol harms in a high-risk group, but only if members of this group are consuming no/lo drinks as a substitute for alcohol.

Existing research on whether people consume no/lo drinks as a substitute for, or in addition to, alcohol is limited. Whilst free provision ^25^ and increased availability of no/lo drinks ^26^ has been shown to reduce alcohol consumption in experimental studies, observational research ^27,28^ shows limited evidence for substitution in, with small reductions in some subgroups but not in others. Self-report data also suggests heterogeneity in this behaviour: a UK survey of past and present no/lo drinkers found that 33% of heavy drinkers consumed no/lo drinks in addition to alcohol, while 25% used them to reduce consumption ^9^.

## CONCLUSION

This study highlights the heterogeneity of low-alcohol and alcohol-free (no/lo) beverage purchasing behaviour in the UK, revealing three distinct household profiles that differ in their purchasing patterns, demographic characteristics, and potential implications for public health. Most no/lo beverage purchasing households purchased no/lo drinks infrequently, limiting the potential for no/lo drinks purchasing to reduce alcohol harms in these households. In contrast, the increased availability of no/lo drinks has the potential to reduce alcohol harms in a smaller but significant group of high-frequency alcohol and no/lo beverage purchasers, but more research is needed to determine whether this group is consuming no/lo beverages in addition to, or as a substitute for, alcohol.

## Data Availability

This study uses Kantar World Panel market research data which cannot be shared by the authors.

## AUTHOR CONTRIBUTIONS

O.Rousham conceptualised the study, carried out analyses, interpretation and writing. A.Stevely and J.Holmes supervised the research, aided with the interpretation of results, and provided feedback on the manuscript.

